# Systemic inflammation and subsequent risk of amyotrophic lateral sclerosis: prospective cohort study

**DOI:** 10.1101/2023.03.06.23286852

**Authors:** G. David Batty, Mika Kivimäki, Philipp Frank, Catharine R. Gale, Liam Wright

## Abstract

**Importance:** While systemic inflammation has been implicated in the aetiology of selected neurodegenerative disorders, its role in the development of amyotrophic lateral sclerosis (ALS) is untested.

**Objective:** To quantify the relationship of C-reactive protein (CRP), an acute-phase reactant and marker of systemic inflammation, with ALS occurrence.

**Design, Setting, Participants:** UK Biobank, a prospective cohort study of 502,649 participants who were aged 37 to 73 years when examined at research centres between 2006 and 2010.

**Exposure:** Venous blood was collected at baseline in the full cohort and assayed for CRP. Repeat measurement was made 3-7 years later in a representative subgroup (N=14,514) enabling correction for regression dilution.

**Main Outcome(s) and Measure(s):** ALS as ascertained via national hospitalisation and mortality registries. We computed multi-variable hazard ratios with accompanying 95% confidence intervals for log-transformed CRP expressed as standard deviation and tertiles.

**Results:** In an analytical sample of 400,884 individuals (218,203 women), a mean follow-up of 12 years gave rise to 231 hospitalisations and 223 deaths ascribed to ALS. After adjustment for covariates which included health behaviours, comorbidity, and socio-economic status, a one standard deviation higher log-CRP was associated with elevated rates of both ALS mortality (hazard ratios; 95% confidence intervals: 1.32; 1.13, 1.53) and hospitalisations (1.20; 1.00, 1.39). There was evidence of dose-response effects across tertiles of CRP for both outcomes (p for trend≤0.05). Correction for regression dilution led to a strengthening of the relationship with CRP for both mortality (1.62; 1.27, 2.08) and hospitalisations (1.37; 1.05, 1.76) ascribed to ALS.

**Conclusions and Relevance:** Higher levels of CRP, a blood-based biomarker widely captured in clinical practice, were associated with a higher subsequent risk of ALS.

**Key Points:** *Question:* Is C-reactive protein (CRP), a marker of systemic inflammation widely used in clinical practice, associated with later risk of amyotrophic lateral sclerosis (ALS)?

*Findings:* Following 11 years disease surveillance in 400,884 individuals (218,203 women), after adjustment for covariates and correction for regression dilution, a one standard deviation higher CRP levels were associations with both mortality (hazard ratio; 95% confidence interval: 1.62; 1.27, 2.08) and hospitalisations (1.37; 1.05, 1.76) ascribed to ALS.

*Meaning:* In the present study, CRP has a dose-response relationship with the risk of later ALS.

## Introduction

Amyotrophic lateral sclerosis (ALS), also known as motor neuron disease, involves the unabated degeneration of nerve cells responsible for voluntary muscle movement. With there being no effective treatment, death from respiratory failure typically occurs within 3 years of symptom emergence.^1,2^ This brings into sharp focus the need for primary prevention research, yet series of studies examining the role of modifiable risk factors, including biomarkers, have revealed disappointing results.^3-6^

ALS is characterised by neuro-inflammation^7^ and there is some evidence implicating C-reactive protein (CRP), an acute-phase reactant and widely used marker of systemic inflammation, in the disease process. In animal models, higher levels of CRP appear to increase the permeability of the blood–brain barrier, so triggering microglial activation^8^ leading to increased release of proinflammatory cytokines, neuroinflammation, and cell death in the brain.^9^ Evidence for a role of CRP in the aetiology of ALS in humans is, however, largely circumstantial. While case-control studies have found that people with established ALS had higher CRP levels relative to the disease-free,^10,11^ this could be a consequence of the condition rather than a cause. Relatedly, surveillance of ALS patients reveals those with elevated systemic inflammation experienced a greater subsequent burden of disease severity, disability, and case fatality.^10,12,13^

The absence of evidence from well-characterised prospective studies on the potential role of CRP as a risk indicator for the onset of ALS is in part ascribed to the rarity of this disorder, rendering datasets insufficiently powered to yield sufficient cases to facilitate analyses. In UK Biobank,^14^ a prospective cohort study, a pre-morbid measurement of CRP was made in a cohort of 0.5 million members of the general population. Importantly, the repeat capturing of this inflammatory marker provides an unusual opportunity to address the impact of well-documented time-dependent fluctuations in CRP when measurement on a single baseline occasion is likely to lead to an underestimation of the true magnitude of the relationship with ALS.^15^

## Methods

UK Biobank is a UK-wide, on-going, closed, prospective cohort study. Described in detail elsewhere,^5^ between 2006 and 2010, 502,649 people aged 37 to 73 years attended 22 geographically disparate research clinics where they completed a questionnaire, underwent an interview, and took part in a medical examination. Ethical approval was obtained from the National Health Service National Research Ethics Service with all participants providing written consent. Using pseudonymised data, the present analyses did not require additional permissions. The present report follows STROBE guidelines for the presentation of original epidemiological research.^16^

### Assessment of baseline characteristics

Ethnicity was self-reported and categorised as White, Asian, Black, Chinese, Mixed, or other ethnic group.^17^ Cigarette smoking and physical activity were measured using standard enquiries.^18^ Self-reported physician diagnosis was collected for ALS, vascular or heart problems, diabetes, and cancer, and study members were asked if they had ever been under the care of a psychiatrist.^19^ The Townsend deprivation index, widely-used as an indicator of neighbourhood socioeconomic circumstances,^20^ is continuously scored with higher values denoting greater deprivation.

Height and weight were measured directly and body mass index was calculated using the usual formulae (weight, kg/height^2^, m^2^).^21^ Forced expiratory volume in one second, a measure of pulmonary function, was quantified using spirometry with the best of three technically satisfactory exhalations used in our analyses. The best result from 3 trials on each hand using a hydraulic hand dynamometer provided a measure of handgrip strength. Non-fasting venous blood was drawn, with assaying conducted at a dedicated central laboratory for CRP, glycated haemoglobin (HbA1c), and high-density lipoprotein (LDL) cholesterol.^22^ At resurvey, 3-7 years after baseline examination, a random sample of baseline study members (N=17,835) had their CRP levels reassessed.

### Ascertainment of amyotrophic lateral sclerosis during follow-up

Study participants were linked to the National Health Service’s (NHS) Central Registry which provided vital status data on study members and, where applicable, cause of death.^23^ Linkage was also made to hospital in-patients records via the NHS Hospital Episode Statistics, a registry of all hospitalisations in the UK.^24^ Using both databases, ALS was denoted by the World Health Organization International Classification of Disease (version 10) code G122.

### Statistical analyses

In all analyses, to address concerns regarding reverse causality – the notion that ALS might influence CRP^25^ rather than the opposite – we excluded 85 people who self-reported ALS at baseline medical examination and/or were hospitalised with the disease before study induction. Additionally, to capture study members with potentially subclinical (undiagnosed) ALS, we left-censored study members such that those who were hospitalised for, or died from, the condition within the first 3 years of baseline were also excluded (N=61). Lastly, study members with a CRP value ≥10.0, indicative of an acute infection, were also excluded.^26^ This resulted in analytical sample of 400,884 (218,203 women) with full data on CRP, covariates, and ALS outcomes.

To summarise the association between CRP and ALS, we used Cox proportional hazards regression to compute hazard ratios with accompanying 95% confidence intervals.^27^ In these analyses, calendar period was the time scale and study members were censored at date of hospitalisation or death from ALS, or end of follow-up (23 March 2021 for mortality, 5 May 2021 for hospitalisation) – whichever came first. CRP was utilised as a standardised, log-transformed linear term (mean=0, SD=1) and as tertiles. To examine potential non-linearity in the association between CRP levels and rates of ALS, we included CRP in models using natural cubic splines. To account for regression dilution bias, we also ran models including a dilution corrected value for CRP following a univariate regression calibration procedure.^15^ This involved regressing the second measure on the first and using the model predictions to calculate corrected CRP values for the full analytic sample. Using this derived value for CRP in the Cox model, confidence intervals were estimated via bootstrapping (1,000 replications).

## Results

In table 1 we show baseline data according to tertiles of CRP. As anticipated, less favourable risk factor levels were universally apparent in people with higher CRP. For instance, people with higher levels of CRP tended to be older, have higher body mass index, lower lung function, and lower socio-economic position. Study members with higher CRP also had a greater prevalence of cigarette smoking, physical inactivity, and an array of morbidities, including vascular disease, cancer, diabetes, and mental illness.

**Table 1.** Baseline characteristics according to C-reactive protein levels in UK Biobank (2006-2020)

|  | **Tertile 1**<br>**(≤0.78 mg/L)** | **Tertile 2**<br>**(0.79 - 1.88 mg/L)** | **Tertile 3**<br>**(1.89 - 9.99 mg/L)** |
| --- | --- | --- | --- |
| Number of people (women) | 134,572 (73,296) | 132,984 (68,479) | 133,328 (76,428) |
| C-Reactive Protein, mg/L | 0.47 (0.18) | 1.27 (0.31) | 3.85 (1.86) |
| Age, yr | 54.99 (8.16) | 56.74 (8.03) | 57.29 (7.92) |
| Glycated haemoglobin, mmol/mol | 34.75 (5.18) | 35.72 (5.94) | 37.09 (7.47) |
| Grip strength, kg | 33.79 (11.18) | 33.38 (11.33) | 31.58 (11.19) |
| High-density lipoprotein cholesterol, mmol/L | 1.55 (0.39) | 1.45 (0.38) | 1.38 (0.35) |
| Forced Expiratory Volume in 1 sec, L | 3.03 (0.8) | 2.88 (0.78) | 2.69 (0.76) |
| Body mass index, kg/m<sup>2</sup> | 24.96 (3.43) | 27.16 (3.89) | 29.45 (5) |
| Neighbourhood deprivation score | -1.58 (2.94) | -1.49 (2.99) | -1.15 (3.14) |
| Non-White, N (%) | 6,767 (5.03) | 6,391 (4.81) | 6,972 (5.23) |
| Current cigarette smoker, N (%) | 10,443 (7.76) | 12,307 (9.25) | 17,028 (12.77) |
| Low physical activity, N (%) | 4,893 (3.64) | 6,843 (5.15) | 11,142 (8.36) |
| Vascular disease, N (%) | 28,251 (20.99) | 37,638 (28.3) | 46,804 (35.1) |
| Diabetes, N (%) | 4,708 (3.5) | 6,039 (4.54) | 8,163 (6.12) |
| Cancer, N (%) | 8,569 (6.37) | 9,527 (7.16) | 10,785 (8.09) |
| Psychiatric consultation, N (%) | 13,769 (10.23) | 13,900 (10.45) | 16,206 (12.15) |

Results are mean (SD) unless otherwise stated.
All baseline characteristics revealed statistical heterogeneity across tertiles of CRP (p-value $p < 0.0001$ )

Mortality surveillance resulted in 223 deaths attributed to ALS, and in these analyses higher baseline CRP levels were related to a higher subsequent risk of this disorder (table 2 and supplemental figure 1). Thus, after adjustment for multiple covariates, the highest tertile of systemic inflammation was associated with a 75% increase in the risk of death from ALS (hazard ratio; 95% confidence interval: 1.75; 1.21, 2.53). A dose-response relationship was also apparent (p-value for trend <0.001) such that the central tertile of CRP had intermediate risk (1.46; 1.02, 2.08). When expressed per standard deviation increase in log of baseline CRP, there was a 32% increase in the risk of ALS (1.32; 1.13, 1.53). When we utilised resurvey data on CRP levels to examine the impact of repression dilution (correlation coefficient for survey-resurvey CRP was 0.64 [N= 14,514, p-value < 0.001; figure 1]), the magnitude of relative risk per SD approximately doubled with this correction (1.62; 1.27, 2.08).

**Table 2.** Association of baseline C-reactive protein (2006-2010) with ALS risk (2010-2020) in UK Biobank (N=400,884)

|  |  | **Tertile 1**<br>**(≤0.78 mg/L)** | **Tertile 2**<br>**(0.79 - 1.88 mg/L)** | **Tertile 3**<br>**(1.89 - 9.99 mg/L)** | **p-value**<br>**for trend** | **Per 1 SD increase**<br>**(observed)** | **Per 1 SD increase**<br>**(dilution corrected)** |
| --- | --- | --- | --- | --- | --- | --- | --- |
| Death | N cases / N at risk | 53 / 134,572 | 80 / 132,984 | 90 / 133,328 |  |  |  |
|  | Age- & sex-adjusted | 1.00 (ref) | 1.35 (0.95, 1.91) | 1.49 (1.06, 2.10) | 0.02 | 1.21 (1.06, 1.39) | 1.38 (1.11, 1.69) |
|  | Multiply-Adjusted | 1.00 (ref) | 1.46 (1.02, 2.08) | 1.75 (1.21, 2.53) | < 0.001 | 1.32 (1.13, 1.53) | 1.62 (1.27, 2.08) |
| Hospitalisation | N cases / N at risk | 62 / 134,572 | 79 / 132,984 | 90 / 133,328 |  |  |  |
|  | Age- & sex-adjusted | 1.00 (ref) | 1.16 (0.83, 1.61) | 1.30 (0.94, 1.80) | 0.11 | 1.14 (0.99, 1.30) | 1.24 (1.00, 1.53) |
|  | Multiply-Adjusted | 1.00 (ref) | 1.22 (0.87, 1.71) | 1.43 (1.00, 2.04) | 0.05 | 1.20 (1.03, 1.39) | 1.37 (1.05, 1.76) |

Range of CRP values according to tertile: 1 (≤0.78); 2 (0.79, 1.88); and 3 (1.89, 9.99 mg/L).
Multiple adjustment is adjustment for: age, sex, ethnicity, smoking status, physical activity, body mass index, co-morbidity (vascular disease, diabetes, cancer, mental illness), lung function, and Townsend deprivation score

**Figure 1.**
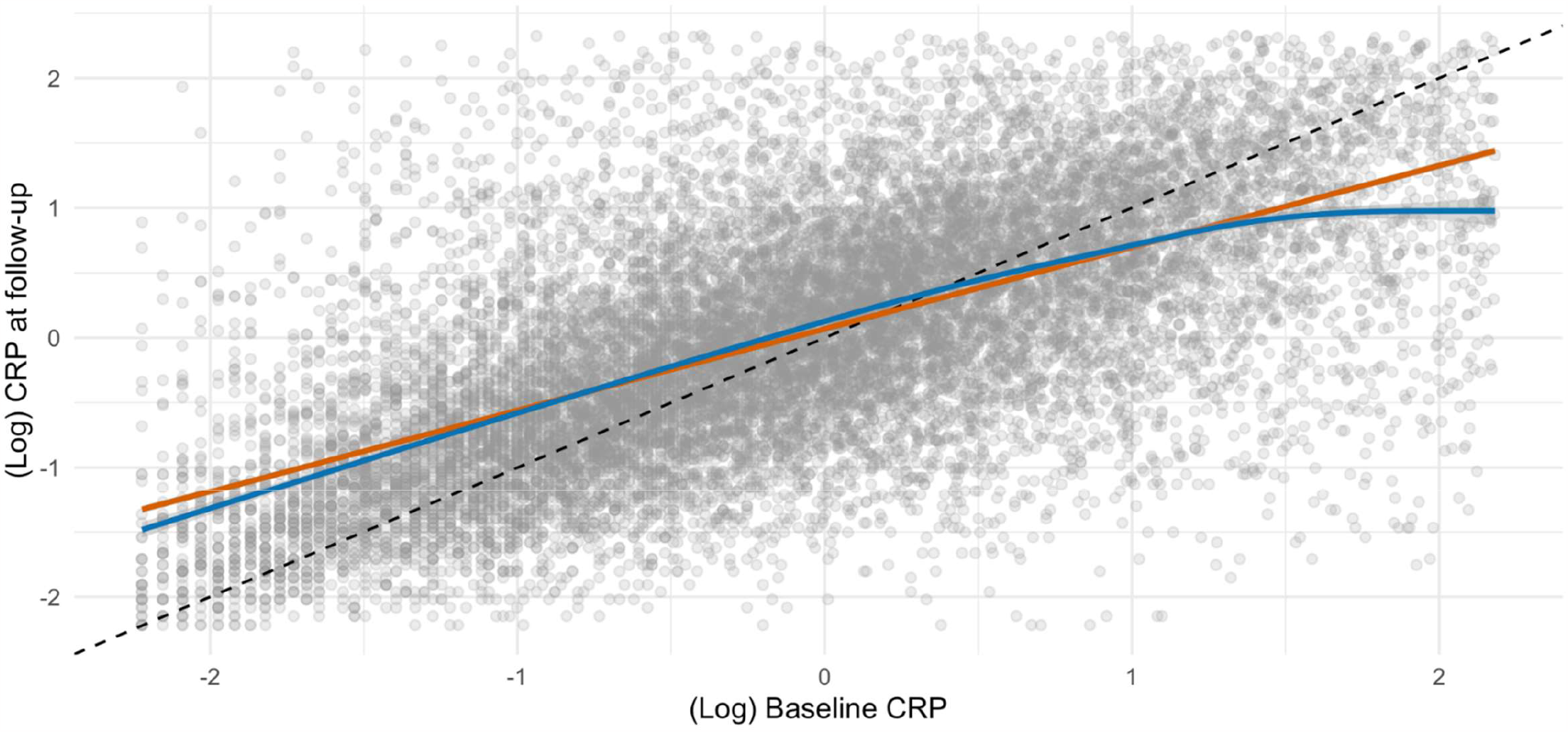
Scatterplot of CRP from baseline and follow-up (N=17,835)

A mean follow-up of 12.0 years (range 0.01-14.4) gave rise to 231 hospitalisations for ALS (117 women). The pattern of association for hospitalisation and CRP was similar to that seen for mortality but effect estimates were typically of lower magnitude and did not always achieve statistical significance at conventional levels (table 2 and supplemental figure 1). Again, a strengthening of the CRP–ALS association was apparent after taking into account regression dilution bias. Thus, the multiply-adjusted hazard ratio per SD increase in CRP increased from 1.20 (1.03, 1.39) to 1.37 (1.05, 1.76).

We also conducted some sensitivity analyses. First, splines allowed us to scrutinise these apparent dose-response effects for CRP in relation to ALS (figure 2) by searching for any inflection that would have been hidden by analyses of the categorisation of CRP. There was a suggestion that the steepest elevation in risk was apparent at lower levels of CRP with a less pronounced increase thereafter. The shape of the association was broadly similar for both deaths and hospitalisations. Second, just as there will be variation in CRP values over time, levels of the covariates in the present study are similarly time-dependent. Correcting for covariate biomarkers (HbA1c, HDL, FEV1 and grip strength), in addition to CRP, revealed very similar results to those apparent when correcting for CRP alone: hospitalisation for ALS (1.37; 1.05, 1.80) and mortality from ALS (1.64; 1.24, 2.10).

**Figure 2.**
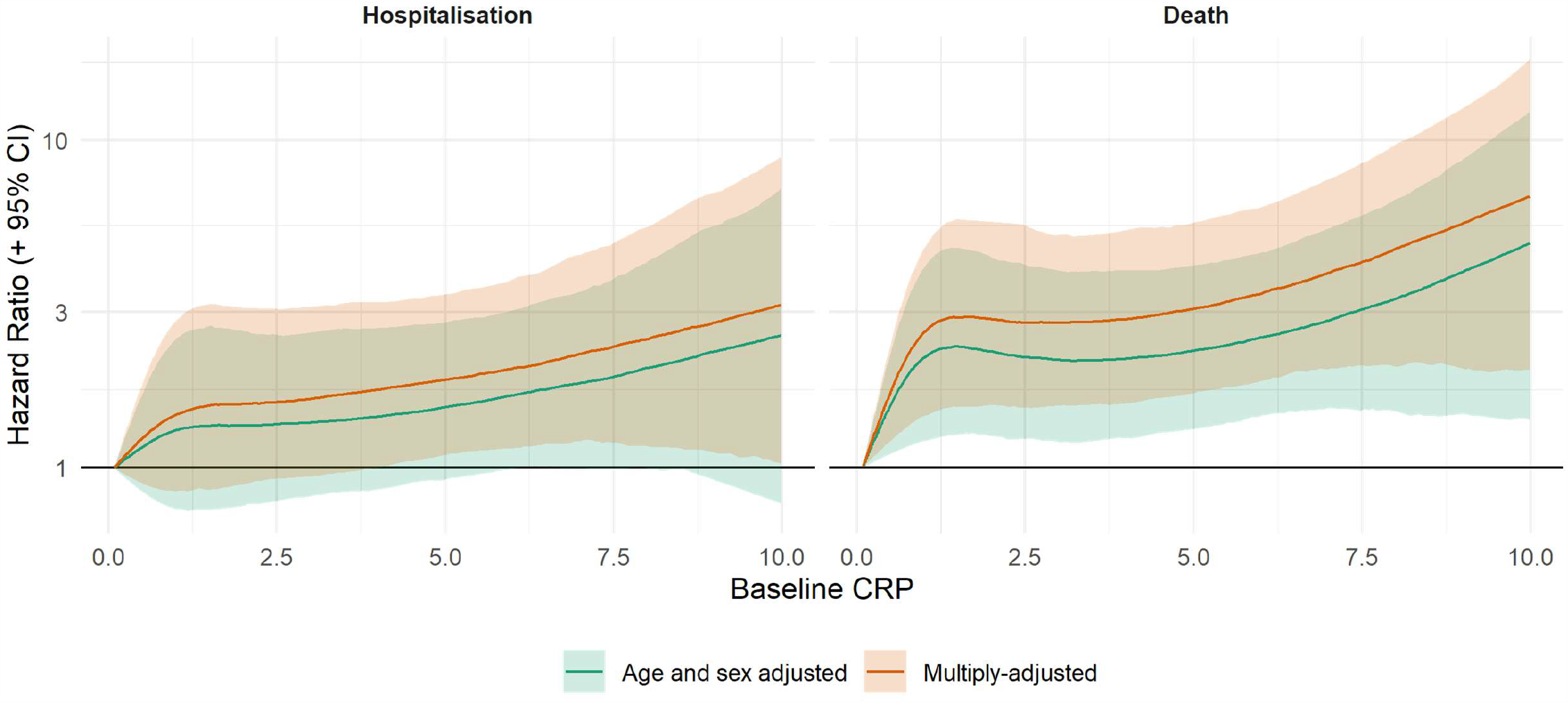
Association of C-reactive protein (2006-2020) with ALS risk (2010-2020) in UK Biobank (N=400,884)

## Discussion

Our main findings were that, in a population initially apparently free of ALS, higher baseline CRP was related to a subsequently elevated risk of developing this neurodegenerative disorder. This association was stronger for ALS mortality relative to hospitalisation; strengthened by adjustment for multiple confounding factors; and of markedly higher magnitude after correction for regression dilution. The two non-genetic factors most closely linked to the development of ALS – age and sex^28^ – were recapitulated here. Thus, people who were older (hazard ratio; 95% confidence interval per decade increase: 2.08; 1.72, 2.50) and male (1.16; 0.90, 1.50) experienced elevated rates of hospitalisation (corresponding results for death from ALS were 1.20; 0.92, 1.56; and 2.42; 1.98, 2.95). This gives us some confidence in the novel results for CRP.

### Comparison with other studies

Elevated levels of systemic inflammation have been shown to be prospectively associated with higher rates of other neurodegenerative disorders such as dementia^29^ and Parkinson’s disease.^30^ With this study being, to our knowledge, the first to specifically examine the role of pre-morbid CRP as a potential risk factor for ALS occurrence, direct comparison of our results with other investigations is not possible. In a study most closely resembling our own, however, haptoglobin, a little-utilised marker of systemic inflammation, was not related to the development of ALS 15 years later.^31^ In vitro and animal work suggesting that the actions of the neuroinflammation-promoting enzyme, cyclooxygenase-2, are inhibited by non-steroidal anti-inflammatory drugs.^32,33^ In a related epidemiological study, however, investigators examining regular use of nonsteroidal anti-inflammatory medication prior to symptom onset found no relationship with ALS. ^34 35^

That the association between CRP and ALS was robust to the adjustment of potential confounding and meditating factors raises the suggestion of a direct effect. The observation that the steepest elevation in risk was apparent at lower levels of CRP with a less pronounced increase thereafter suggests the mechanism relates to low-grade systemic inflammation. One possibility is that even modestly raised of CRP increase the permeability of the blood–brain barrier, so triggering microglial activation^8^ leading to enhanced release of proinflammatory cytokines, neuroinflammation, and cell death in the brain.^36^ It was not possible to directly explore these potential explanations using the present data, however.

### Study strengths and limitations

The strengths of the present study include its novelty; the use of recapture data on CRP to examine regression dilution bias; its scale, which facilitates the accumulation of a sufficiently high number of cases for analyses of a rare neurodegenerative disorder alongside left-censoring to take into account reverse causality; and the well-characterised nature of the study participants which facilitates adjustment for multiple confounding factors.

Inevitably, however, our work has its weaknesses. First, the present study sample comprises only the 5.5% of the target population.^14^ As has been demonstrated,^37,38^ the data material is therefore inappropriate for estimation of risk factor or disease prevalence and for event incidence, including for ALS. These observations do not, however, seem to influence reproducibility of the association of established risk factors for important health outcomes such as vascular disease, selected cancers, and suicide.^38^ We think the same reasoning can be applied to the present analyses for ALS. Second, we did not have data on other markers of systemic inflammation such as interleukin 6 with which to draw comparison with the present results for CRP. Lastly, the outcomes herein represent advanced ALS and it is unknown if CRP is related to ALS at earlier stages of disease progression.

In conclusion, in the present study, higher levels of pre-morbid CRP, an inflammatory marker commonly captured in clinical practice, appeared to be associated with higher risk of incident ALS. These results warrant further research examining the mechanisms linking systemic inflammation to ALS pathology.

## Data Availability

Data from UK Biobank upon application (http://www.ukbiobank.ac.uk/). Part of the present research has been conducted using the UK Biobank Resource under Application 10279.

## Supplemental figures

**Supplemental figure 1.**
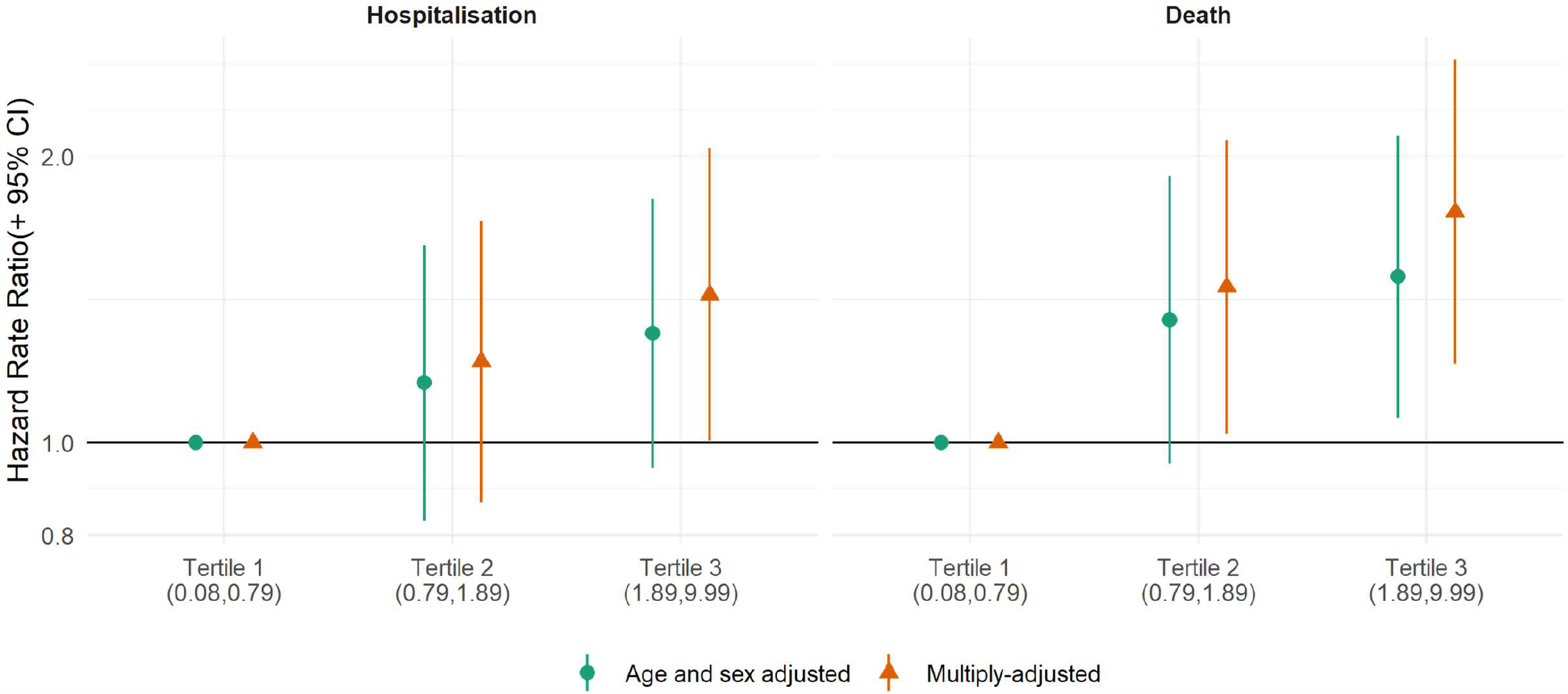
Association of C-reactive protein (2006-2010) with ALS risk (2010-2020) in UK Biobank (N=400,884)

**Supplemental figure 2.**
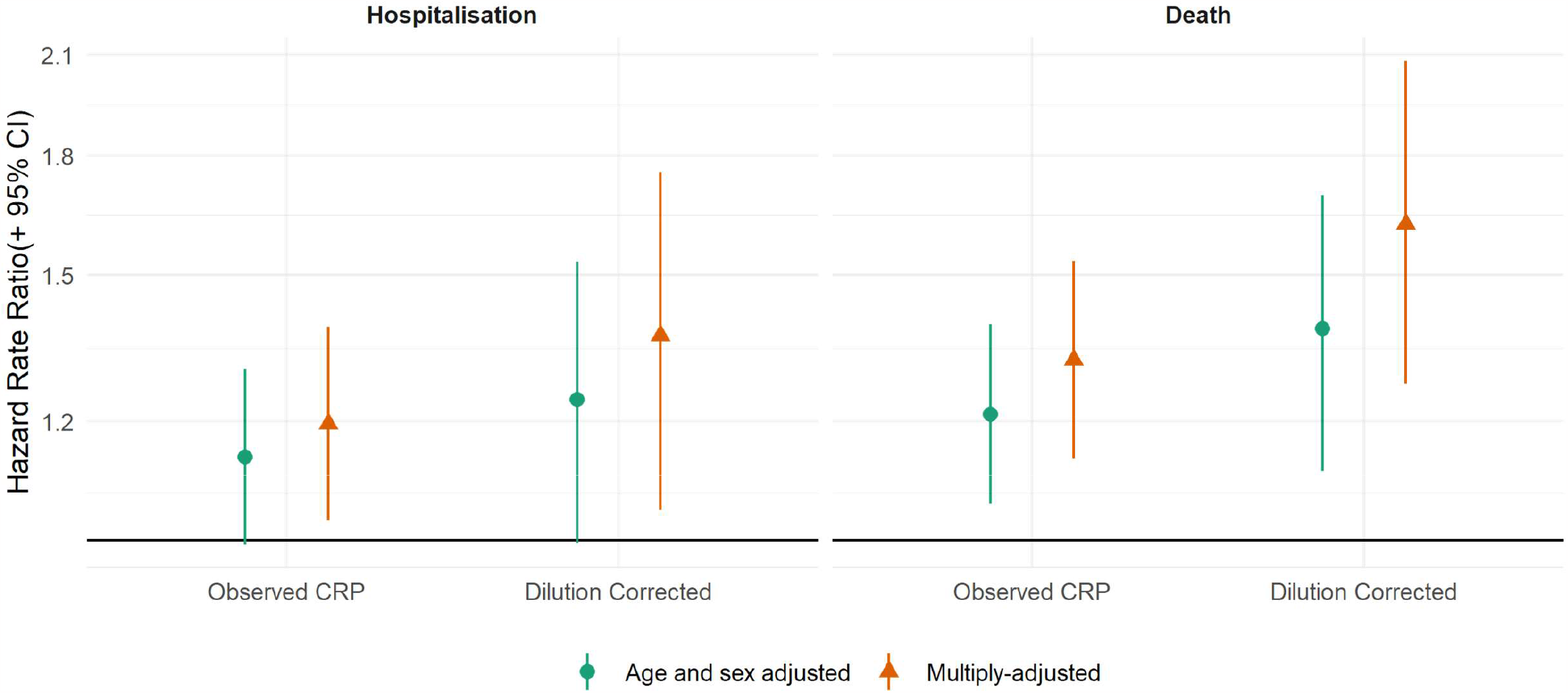
Association of C-reactive protein (2006-2010) with ALS risk (2010-2020) in UK Biobank (N=400,884) – without and with correction for regression dilution bias.

## References

1. Byrne S, Elamin M, Bede P, et al. Cognitive and clinical characteristics of patients with amyotrophic lateral sclerosis carrying a C9orf72 repeat expansion: a population-based cohort study. Lancet Neurol. Mar 2012;11(3):232–40. doi:10.1016/s1474-4422(12)70014-5

2. Abhinav K, Stanton B, Johnston C, et al. Amyotrophic lateral sclerosis in South-East England: a population-based study. The South-East England register for amyotrophic lateral sclerosis (SEALS Registry). Neuroepidemiology. 2007;29(1-2):44–8. doi:10.1159/000108917

3. Ingre C, Roos PM, Piehl F, Kamel F, Fang F. Risk factors for amyotrophic lateral sclerosis. Clin Epidemiol. 2015;7:181–93. doi:10.2147/clep.S37505

4. Al-Chalabi A, Hardiman O. The epidemiology of ALS: a conspiracy of genes, environment and time. Nat Rev Neurol. Nov 2013;9(11):617–28. doi:10.1038/nrneurol.2013.203

5. Couratier P, Corcia P, Lautrette G, Nicol M, Preux PM, Marin B. Epidemiology of amyotrophic lateral sclerosis: A review of literature. Rev Neurol (Paris). Jan 2016;172(1):37–45. doi:10.1016/j.neurol.2015.11.002

6. Oskarsson B, Gendron TF, Staff NP. Amyotrophic Lateral Sclerosis: An Update for 2018. Mayo Clin Proc. Nov 2018;93(11):1617–1628. doi:10.1016/j.mayocp.2018.04.007

7. Philips T, Robberecht W. Neuroinflammation in amyotrophic lateral sclerosis: role of glial activation in motor neuron disease. Lancet Neurol. Mar 2011;10(3):253–63. doi:10.1016/s1474-4422(11)70015-1

8. Hsuchou H, Kastin AJ, Mishra PK, Pan W. C-reactive protein increases BBB permeability: implications for obesity and neuroinflammation. Cell Physiol Biochem. 2012;30(5):1109–19. doi:10.1159/000343302

9. Beers DR, Zhao W, Neal DW, et al. Elevated acute phase proteins reflect peripheral inflammation and disease severity in patients with amyotrophic lateral sclerosis. Sci Rep. Sep 17 2020;10(1):15295. doi:10.1038/s41598-020-72247-5

10. Keizman D, Rogowski O, Berliner S, et al. Low-grade systemic inflammation in patients with amyotrophic lateral sclerosis. Acta Neurol Scand. Jun 2009;119(6):383–9. doi:10.1111/j.1600-0404.2008.01112.x

11. Ryberg H, An J, Darko S, et al. Discovery and verification of amyotrophic lateral sclerosis biomarkers by proteomics. Muscle Nerve. Jul 2010;42(1):104–11. doi:10.1002/mus.21683

12. Sun J, Carrero JJ, Zagai U, et al. Blood biomarkers and prognosis of amyotrophic lateral sclerosis. Eur J Neurol. Nov 2020;27(11):2125–2133. doi:10.1111/ene.14409

13. Lunetta C, Lizio A, Maestri E, et al. Serum C-Reactive Protein as a Prognostic Biomarker in Amyotrophic Lateral Sclerosis. JAMA Neurol. Jun 1 2017;74(6):660–667. doi:10.1001/jamaneurol.2016.6179

14. Sudlow C, Gallacher J, Allen N, et al. UK biobank: an open access resource for identifying the causes of a wide range of complex diseases of middle and old age. PLoS Med. 3/2015 2015;12(3):e1001779. Not in File.

15. Keogh RH, White IR. A toolkit for measurement error correction, with a focus on nutritional epidemiology. Stat Med. May 30 2014;33(12):2137–55. doi:10.1002/sim.6095

16. von Elm E, Altman DG, Egger M, et al. The Strengthening the Reporting of Observational Studies in Epidemiology (STROBE) statement: guidelines for reporting observational studies. Lancet. Oct 20 2007;370(9596):1453–7. doi:10.1016/S0140-6736(07)61602-X

17. Lassale C, Gaye B, Hamer M, Gale CR, Batty GD. Ethnic disparities in hospitalisation for COVID-19 in England: The role of socioeconomic factors, mental health, and inflammatory and pro-inflammatory factors in a community-based cohort study. Brain Behav Immun. Aug 2020;88:44–49. doi:10.1016/j.bbi.2020.05.074

18. Hamer M, Kivimaki M, Gale CR, Batty GD. Lifestyle risk factors, inflammatory mechanisms, and COVID-19 hospitalization: A community-based cohort study of 387,109 adults in UK. Brain Behav Immun. Jul 2020;87:184–187. doi:10.1016/j.bbi.2020.05.059

19. Batty GD, McIntosh AM, Russ TC, Deary IJ, Gale CR. Psychological distress, neuroticism, and cause-specific mortality: early prospective evidence from UK Biobank. J Epidemiol Community Health. 11/2016 2016;70(11):1136-1139. Not in File.

20. Batty GD, Deary IJ, Luciano M, Altschul DM, Kivimaki M, Gale CR. Psychosocial factors and hospitalisations for COVID-19: Prospective cohort study based on a community sample. Brain Behav Immun. Oct 2020;89:569–578. doi:10.1016/j.bbi.2020.06.021

21. Hamer M, Gale CR, Kivimaki M, Batty GD. Overweight, obesity, and risk of hospitalization for COVID-19: A community-based cohort study of adults in the United Kingdom. Proc Natl Acad Sci U S A. Sep 1 2020;117(35):21011–21013. doi:10.1073/pnas.2011086117

22. Hamer M, Gale CR, Batty GD. Diabetes, glycaemic control, and risk of COVID-19 hospitalisation: Population-based, prospective cohort study. Metabolism. Aug 22 2020;112:154344. doi:10.1016/j.metabol.2020.154344

23. Batty GD, Gale CR, Kivimaki M, Bell S. Assessment of Relative Utility of Underlying vs Contributory Causes of Death. JAMA Open Network. 2019;

24. Abell JG, Lassale C, Batty GD, Zaninotto P. Risk Factors for Hospital Admission After a Fall: A Prospective Cohort Study of Community-Dwelling Older People. J Gerontol A Biol Sci Med Sci. Mar 31 2021;76(4):666–674. doi:10.1093/gerona/glaa255

25. Cui C, Sun J, Pawitan Y, et al. Creatinine and C-reactive protein in amyotrophic lateral sclerosis, multiple sclerosis and Parkinson’s disease. Brain Commun. 2020;2(2):fcaa152. doi:10.1093/braincomms/fcaa152

26. Akbaraly TN, Hamer M, Ferrie JE, et al. Chronic inflammation as a determinant of future aging phenotypes. CMAJ. 11/5/2013 2013;185(16):E763–E770. Not in File. doi:cmaj.122072 [pii];10.1503/cmaj.122072 [doi]

27. Cox DR. Regression models and life-tables. J R Stat Soc [Ser B]. 1972 1972;34:187-220. Not in File.

28. Armon C. An evidence-based medicine approach to the evaluation of the role of exogenous risk factors in sporadic amyotrophic lateral sclerosis. Neuroepidemiology. Jul-Aug 2003;22(4):217–28. doi:10.1159/000070562

29. Kuo HK, Yen CJ, Chang CH, Kuo CK, Chen JH, Sorond F. Relation of C-reactive protein to stroke, cognitive disorders, and depression in the general population: systematic review and meta-analysis. Lancet Neurol. Jun 2005;4(6):371–80. doi:10.1016/S1474-4422(05)70099-5

30. Qiu X, Xiao Y, Wu J, Gan L, Huang Y, Wang J. C-Reactive Protein and Risk of Parkinson’s Disease: A Systematic Review and Meta-Analysis. Front Neurol. 2019;10:384. doi:10.3389/fneur.2019.00384

31. Yazdani S, Mariosa D, Hammar N, et al. Peripheral immune biomarkers and neurodegenerative diseases: A prospective cohort study with 20 years of follow-up. Ann Neurol. Dec 2019;86(6):913–926. doi:10.1002/ana.25614

32. Carty ML, Wixey JA, Reinebrant HE, Gobe G, Colditz PB, Buller KM. Ibuprofen inhibits neuroinflammation and attenuates white matter damage following hypoxia-ischemia in the immature rodent brain. Brain Res. Jul 21 2011;1402:9–19. doi:10.1016/j.brainres.2011.06.001

33. Klegeris A, McGeer PL. Cyclooxygenase and 5-lipoxygenase inhibitors protect against mononuclear phagocyte neurotoxicity. Neurobiol Aging. Sep-Oct 2002;23(5):787–94. doi:10.1016/s0197-4580(02)00021-0

34. Popat RA, Tanner CM, van den Eeden SK, et al. Effect of non-steroidal anti-inflammatory medications on the risk of amyotrophic lateral sclerosis. Amyotroph Lateral Scler. Jun 2007;8(3):157–63. doi:10.1080/17482960601179456

35. Fondell E, O’Reilly EJ, Fitzgerald KC, et al. Non-steroidal anti-inflammatory drugs and amyotrophic lateral sclerosis: results from five prospective cohort studies. Amyotroph Lateral Scler. Oct 2012;13(6):573–9. doi:10.3109/17482968.2012.703209

36. Beers DR, Appel SH. Immune dysregulation in amyotrophic lateral sclerosis: mechanisms and emerging therapies. Lancet Neurol. Feb 2019;18(2):211–220. doi:10.1016/s1474-4422(18)30394-6

37. Fry A, Littlejohns TJ, Sudlow C, et al. Comparison of Sociodemographic and Health-Related Characteristics of UK Biobank Participants With Those of the General Population. Am J Epidemiol. 11/1/2017 2017;186(9):1026-1034. Not in File.

38. Batty GD, Gale CR, Kivimäki M, Deary IJ, Bell S. Comparison of risk factor associations in UK Biobank against representative, general population based studies with conventional response rates: prospective cohort study and individual participant meta-analysis. BMJ (Clinical research ed). 2020;368

